# A predictive model of a growing fetus

**DOI:** 10.1101/2022.12.22.22283844

**Authors:** Chandrani Kumari, Gautam I Menon, Leelavati Narlikar, Uma Ram, Rahul Siddharthan

## Abstract

Fetal growth is monitored periodically during pregnancy via ultrasound measurements of fetal dimensions such as femur length (FL), head circumference (HC), abdominal circumference (AC), and biparietal diameter (BPD). Multiple growth standards have been published for each of these, which are clinically used to place a fetus on a “growth chart”. These consist of percentile tables varying by weeks of gestation, computed from cohorts of “low-risk” women with healthy lifestyles, living conditions, and clinical parameters. Such charts are prescriptive of ideal growth, but not necessarily descriptive of diverse real-world populations where they may be used. Moreover, they are constructed by pooling all fetal measurements across the cohort, not based on a growth model, and therefore not necessarily predictive of growth of an individual fetus.

We show that the Gompertz model, a standard model for constrained growth, with just three intuitive parameters, convincingly fits the growth of fetal ultrasound biometries. Two of these parameters—*t*_0_ (the inflection time) and *c* (the rate of decrease of growth rate)—can be treated as universal to all fetuses, while the third parameter *A* can be modeled as an overall scale parameter specific to each fetus, which captures the individual variation in growth. On our cohort of 817 pregnant women (“Seethapathy cohort”), we show that not only can the value of *A* for each fetus be inferred from ultrasound data available by the second or the third trimester, but the weight of the baby at delivery can also be predicted with remarkable accuracy using these inferred Gompertz parameters. A model trained on the Seethapathy cohort performs well in estimating the birth weight in an independent validation cohort of 365 women, demonstrating the predictive power of the model. Moreover, we find that deviation from Gompertz-like growth is linked to neonatal complications. Finally, we show that the Gompertz growth curve is a close fit to the standards from WHO, NICHD and INTERGROWTH, with the optimal *t*_0_ and *c* close to that in the Seethapathy cohort. We propose that the Gompertz formula be a basis for future growth standards, with almost all variation described by a single scale parameter *A*, which can serve either as a descriptor of mean or variance in population, or as a descriptor for growth of an individual fetus. Indeed, the formula is descriptive of typical growth, predictive of future growth, and may be used in prescriptive standards.

## Introduction

Assessment of fetal growth is an integral part of antenatal care. Babies that are born small or large for their gestational age are known to be at an increased risk for adverse pregnancy outcomes. Fetuses that are small for gestational age (SGA) have an increased risk of stillbirth, neonatal death, neurodevelopmental delay and poor school performance, obesity in childhood and adulthood, and metabolic disease [1]. Being large for gestational age (LGA) is associated with traumatic-composite neonatal morbidity [2], and may cause longer and problematic labor as a result of their physical size. Risks include birth damage, the need for surgical vaginal delivery, or cesarean section. Perinatal morbidity in LGA neonates is associated with postpartum hemorrhage, shoulder dystocia, neonatal hypoglycemia, hyperbilirubinemia, and respiratory complications [3, 4].

The growth of a human embryo is monitored by ultrasound biometric measurements, most commonly of femur length (FL), abdominal circumference (AC), head circumference (HC) and biparietal diameter (BPD). The growth curve of each of these has a sigmoidal shape, with the rate of growth larger midway through gestation and smaller in early and late pregnancy. Ideal growth is described in tabular and graphical form in publications from WHO [5, 6], NICHD [7, 8] and others. Various formulas have been proposed to fit the empirical growth curve. Todros *et al*. (1987) [9] proposed and evaluated three sigmoidal formulas: a cubic polynomial function *a* + *bt* + *ct*^3^, a logistic-logarithmic function *a/*(1 + *bt* − *c*), and an exponential-power function *at*^2^*e*^−*bt*^ (where *t* is the menstrual or the corrected age, and *a, b, c* are parameters). The INTERGROWTH consortium [10] proposed growth standards for standard ultrasound biometries, in the form of formulas derived from regression fitting to a large dataset.

Wosilait *et al*. (1992) [11] proposed using the Gompertz growth formula, which we parametrize as *f* (*t*) = *A* exp (−exp (−*c*(*t* − *t*_0_))) (see Methods for a derivation and explanation of the parametrization), to describe the growth of fetal *volume*. The formula was originally proposed by Gompertz [12], in the context of modeling population size as a function of age. It has since been widely employed in population dynamics, in many other systems including tumor growth [13], and has been argued to be widely applicable to cell population growth [14]. Tjørve and Tjørve [15] provide a detailed review of uses of this model and its variants in growth analysis.

Wosilait *et al*. used pooled fetal data from various sources to show that the Gompertz equation accurately modeled the growth of fetal volume from 50 days post-conception to term. Their data was obtained from four principal sources for fetal weight and age, dating back to 1909. In a follow-up paper [16] they improved the fit for the period before 50 days, including a fifth dataset of early (days 4.5 to 17) fetal data. They also motivated the Gompertz formula by arguing that, of three categories of cells—post-mitotic, renewal, and expanding—only the third contributes to growth, and the fraction of expanding cells decreases with time. While they model volume (as an ellipsoid), they note that the specific gravity of the fetus remains almost constant through gestation, increasing slightly from 1.0 to about 1.05, so the results may be converted to fetal weight.

Fetal weight cannot be directly measured during gestation, but an accurate indirect assessment would be valuable in the process of antenatal care. Several attempts have been made to predict fetal weight at term, and estimate fetal weight during gestation, based on ultrasound measurements. Some widely used formulas are due to Shepard et al (1982) [17], Hadlock *et al*. (1985) [18] (who supplied four formulas using different combinations of biometries), and the latest INTERGROWTH consortium, which has conducted the largest collaborative study of fetal and newborn growth till date (2017) [19]. Milner*et al*. (2018) [20], in a systematic review, assessed 11 different formulas for the birth weight estimation and concluded that the third Hadlock formula (using HC, AC and FL) gave the most accurate results with the lowest random errors. They considered only studies where women had their last scans within seven days of delivery. Kong *et al*. (2019) [21] compared the accuracy of the INTERGROWTH formula for estimated birth weight to that of the fourth Hadlock formula (using HC, AC, FL, and BPD) and Shepard formula, again measured within a week before delivery. They noticed that Hadlock had the lowest mean absolute percentage error (MAPE) of the three formulas. Hiwale *et al*. (2019) [22] used multiple stepwise regression and the LASSO regression method to estimate birth weight in the Indian population. They too used ultrasound scans within seven days of delivery in their model, which they reported outperformed literature models in their cohort.

Recently, more advanced machine learning approaches have been also applied to estimate birth weight. Tao *et al*. (2021) [23] proposed a hybrid long-short-term memory (LSTM) technique to predict birth weight for time series data at delivery using maternal parameters (height, weight, age, uterine height, and abdominal circumference of pregnant women), fetal parameters (HC, AC, BPD, FL, and amniotic fluid index of the fetus), and maternal weight increase at six different gestational time points. The hybrid-LSTM indeed outperformed other ML techniques, but its application is confined to clinics that collect such maternal data at multiple time-points, something that is not done routinely.

In this paper, we fit the Gompertz growth formula to our in-house ultrasound data of 817 pregnant women (which we call the “Seethapathy cohort”). While Wosilait *et al*. [11, 16] *used the Gompertz equation for modeling fetal volume*, we use it for the *linear dimensions* of the fetus, which can be measured via ultrasound: in particular HC, AC, BPD, and FL. We note that if linear dimensions of an ellipsoid are described by the Gompertz formula, then so is the volume, with only the parameters *A* and *t*_0_ changing. Moreover, in contrast to Wosilait *et al*. who pool diverse datasets, we show that each fetus can in fact be individually modeled using a fetus-specific scale factor *A*, while the other parameters *t* and *c* of the equation can be modeled globally.

We show that we can “learn” *A* with good accuracy from the early 24 weeks of ultrasound measurements and use this, to predict the finally measured ultrasound parameters, and the fetal birth weight, with high accuracy (mean absolute percentage error in birth weight prediction of 10.1%). This improves to 7.8% if we use all available ultrasounds up until 35 weeks, and is significantly superior to estimated fetal weight formulas in the literature.

We also examine fetuses whose growth differs significantly from the Gompertz formula. We show that growth that deviates from the Gompertz curve is significantly associated with neonatal complications.

Not only does the Gompertz formula have easily interpretable parameters, we argue that growth standards can be framed to use such intuitive formulas rather than computer-generated expressions. We demonstrate that the Gompertz curve in fact fits the WHO, NICHD, and INTERGROWTH growth standards well, with universal *t*_0_ and *c* for each biometry, and the single parameter *A* sufficient to describe variation in fetal growth. Going beyond growth standards that describe optimal growth, our method of using the Gompertz formula offers the researcher flexibility to model the growth of any specific population, or any specific individual fetus, while lending predictive power for birth outcomes based on ultrasound data.

## Materials and Methods

### Study Design and Participants

We study and model the growth of a fetus during pregnancy, using standard ultrasound fetal biometry measurements. We used a dataset of 3,639 pregnant women from Chennai who were registered at Seethapathy Clinic and Hospital, Chennai, between 2015 and 2017 and gave birth there, for whom ultrasound biometries were available. Four ultrasound fetal measurements—the HC, AC, BPD, and FL were considered. Subjects were excluded from the study if they had fewer than three sets of ultrasound measurements or if their first ultrasound was taken after 15 weeks. Only women with singleton pregnancies were considered. With these criteria there were 1,166 eligible subjects.After applying these filters and quality control on the remaining samples, the cohort consisted of 817 pregnant women. For birth weight prediction we discarded subjects whose second ultrasound was done after 24 weeks since a minimum of two measurements were required to fit *A* at 24 weeks. We also omitted subjects whose last ultrasound measurements were before 27 weeks. The resulting cohort for birth weight prediction consisted of 774 women.

Table 1 shows cohort characteristics, which were optionally used in birth weight predictions (supporting figure S1).

**Table 1.** Data summary considered for the study. Parameters measured on first hospital visit except as indicated

| Cohort characteristics<br>($n = 817$) | Mean ± std or % |
| --- | --- |
| Age (years) | $28.1 \pm 3.8$ |
| Height (cm) | $158.6 \pm 5.9$ |
| Weight (kg) | $64.7 \pm 12$ |
| Body mass index ($\text{kg}/\text{m}^2$) | $25.6 \pm 4.4$ |
| Weight gain (kg, from first to last visit) | $9.8 \pm 4.3$ |
| Fasting blood sugar (mg/dL) | $81 \pm 8$ |
| At risk for gestational diabetes mellitus | 50.2% |
| Diagnosed as anemic | 14.9% |
| Diagnosed with hypothyroidism | 15.8% |
| Diagnosed with hypertensive disorders | 2.4% |
| Diagnosed with gestational diabetes mellitus | 22.5% |
| Had neonatal complications | 18.8% |

Neonatal complications, as diagnosed by the hospital, include preterm birth, intra-uterine growth restriction(IUGR)/small for gestational age (SGA), large for gestational age (LGA), sepsis, respiratory distress, hypoglycemia, and one case of intra-uterine fetal death, as well as a small number of other complications including meconium gastritis, urinary tract infection, neonatal hemolytic anemia. Phototherapy was excluded from complications.

### Published growth standards for fetal biometries

The INTERGROWTH consortium [10] has supplied growth standards for five ultrasound biometries, of which we use only FL, AC, HC and BPD. They supply standards for mean and standard deviation of these biometries, which we list in table 2. These formulas differ for each biometry and are not motivated by a growth model. However, visually, all parameters seem to follow a similar sigmoidal growth pattern, of initial slow growth, followed by more rapid growth whose rate decreases after around 30 weeks (cf figure 3 of INTERGROWTH paper [10] and figures 2C and 6B of this paper).

**Table 2.** INTERGROWTH formulas for the mean and standard deviation of various ultrasound biometry parameters

| Ultrasound Measurements |  | INTERGROWTH formula |
| --- | --- | --- |
| Head circumference | Mean | $28.2849 + 1.69267 \times GA^2 - 0.397485 \times GA^2 \times \log(GA)$ |
| | SD | $1.98735 + 0.0136772 \times GA^3 - 0.00726264 \times GA^3 \log(GA) + 0.000976253 \times GA^3 \times (\log(GA))^2$ |
| Biparietal diameter | Mean | $5.60878 + 0.158369 \times GA^2 - 0.00256379 \times GA^3$ |
| | SD | $\exp(0.101242 + 0.00150557 \times GA^3 - 0.000771535 \times GA^3 \log(GA) + 0.0000999638 \times GA^3 \times (\log(GA))^2)$ |
| Abdominal circumference | Mean | $-81.3243 + 11.6772 \times GA - 0.000561865 \times GA^3$ |
| | SD | $-4.36302 + 0.121445 \times GA^2 - 0.0130256 \times GA^3 + 0.00282143 \times GA^3 \log(GA)$ |
| Femur length | Mean | $-39.9616 + 4.32298 \times GA - 0.0380156 \times GA^2$ |
| | SD | $\exp(0.605843 - 42.0014 \times GA^{-2} + 0.00000917972 \times GA^3)$ |

**Fig 1.**
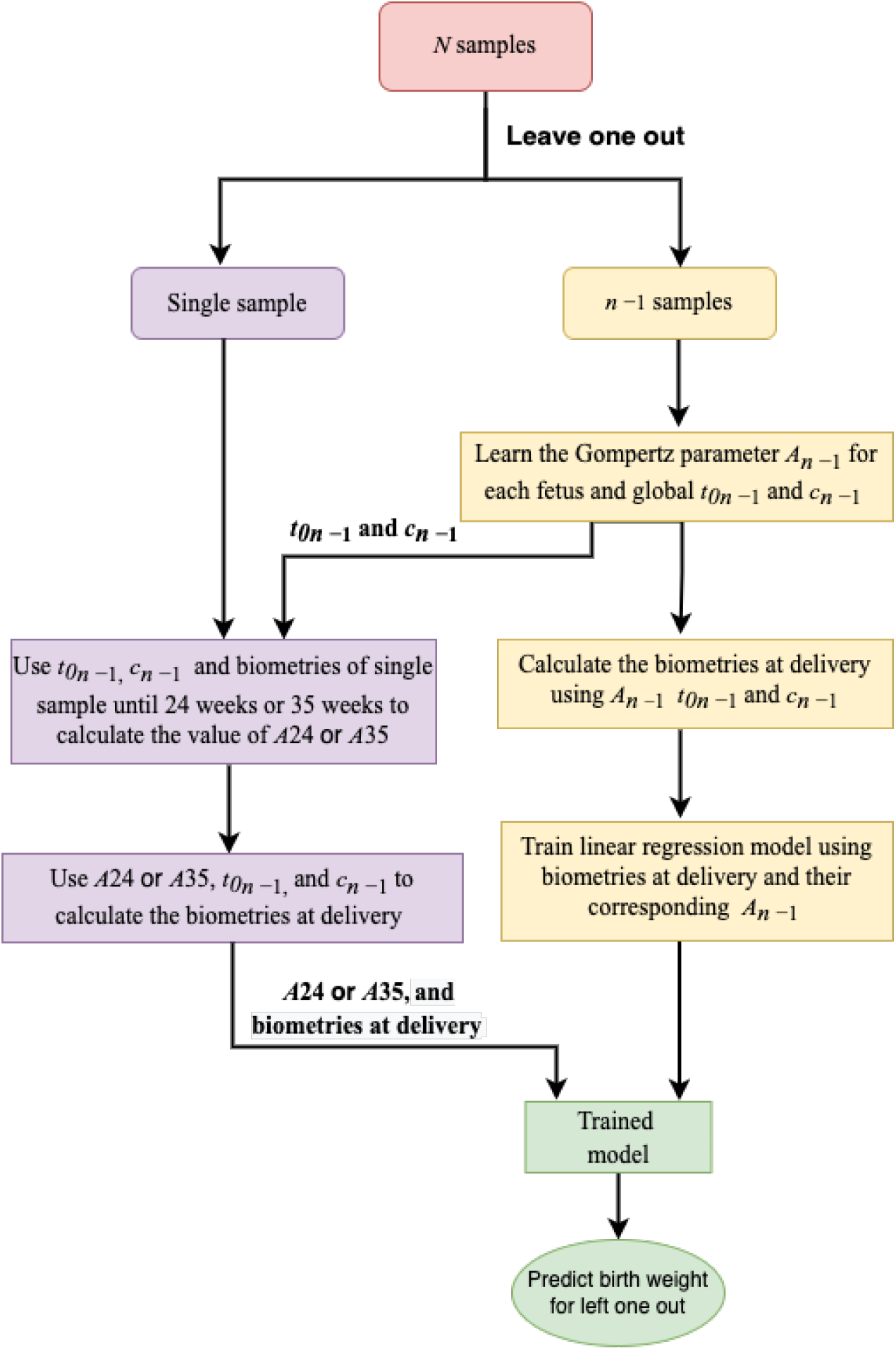
Flowchart for birth weight prediction using a “leave one out” method

**Fig 2.**
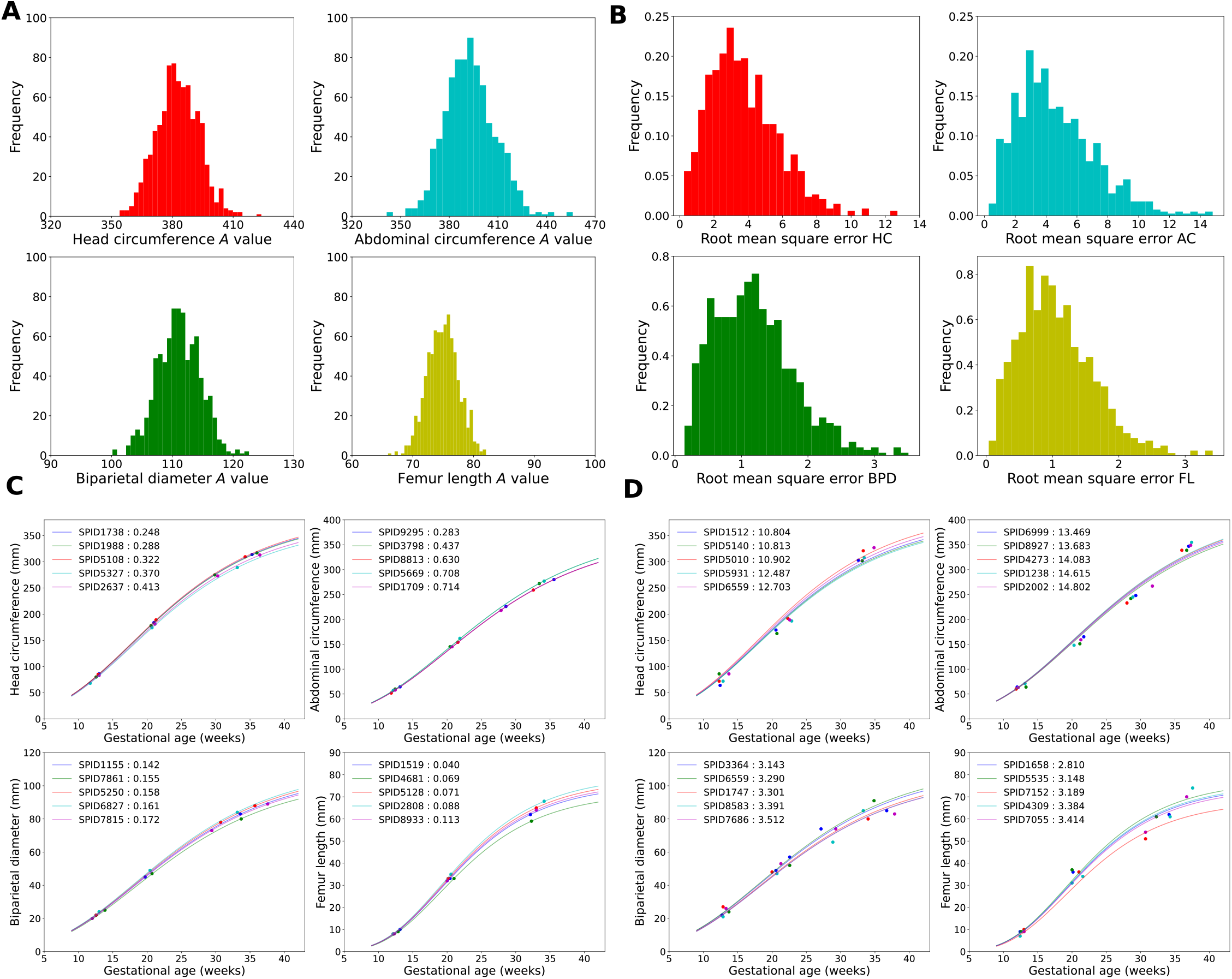
Distribution of *A*, root mean square error and quality of fit to Gompertz equation, **A:** Distribution of *A* for the four biometric parameters we study. **B:** Distribution of the mean squared error for the best fit to the Gompertz function. **C**: Five best-fitting fetuses to the Gompertz curve, for each biometric parameter. **D**: Five worst-fitting fetuses to the Gompertz curve, for each parameter.

**Fig 3.**
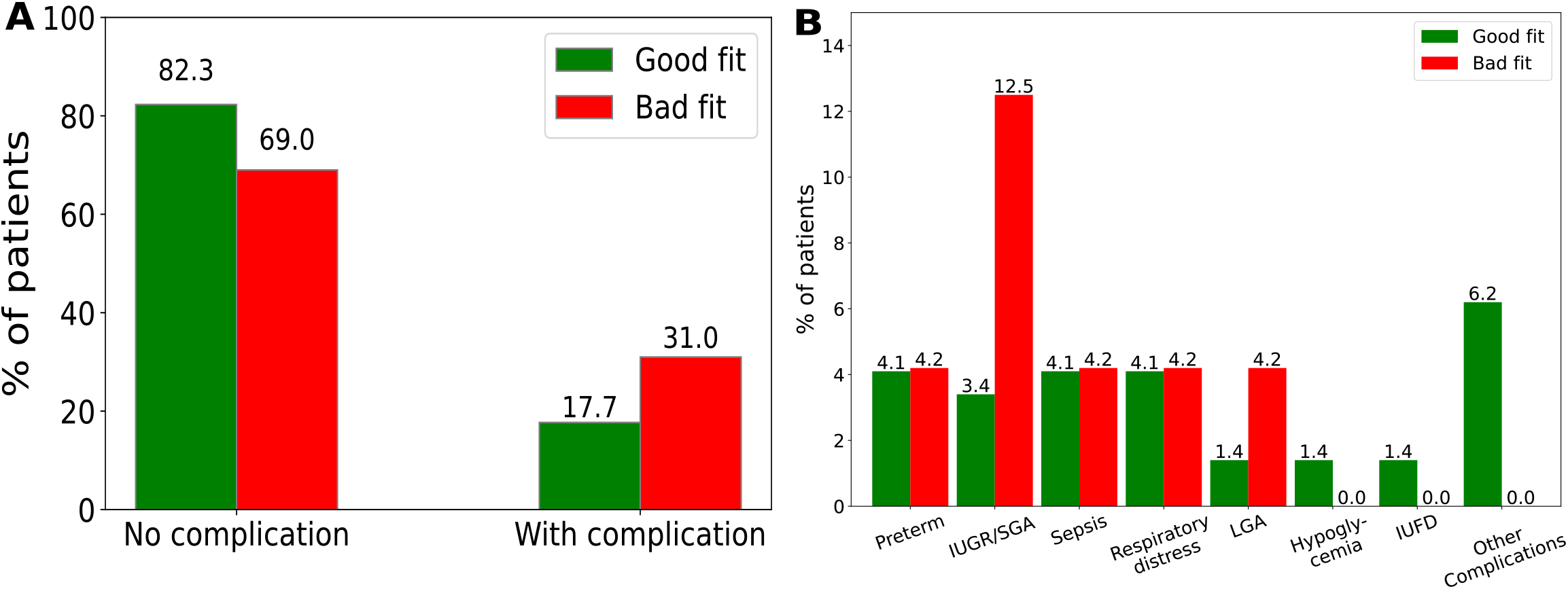
Quality of fit and neonatal complications. **A:** The “overall bad fit” fetuses exhibit a significantly (p = 0.007, hypergeometric test) larger number of neonatal contributions. The percentage of fetuses having neonatal complications is higher in the “overall bad fit” category (31%) than in the “overall good fit” category (17.7%) **B:** The percentage of fetuses having SGA and LGA complications are higher in “overall bad fit” category (12.5% and 4.2% respectively) as compared to “overall good fit” (3.4% and 1.4% respectively).

More recently, the World Health Organization has published fetal growth charts [5], and tables documenting fetal growth of these parameters at various percentiles. We make use of these published INTERGROWTH formulas and WHO tables in comparing with the Gompertz model. A third set of fetal growth charts, which are race-specific and which we do not use, is published by NICHD [7].

### Gompertz growth model

The Gompertz equation for the growth of a parameter *X* as a function of time *t* is:

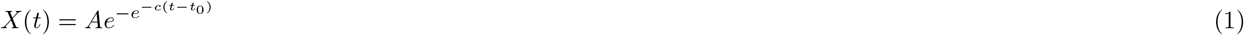

The formula may be viewed as describing a constrained growing system where a parameter *X*(*t*) grows at a rate *r*(*t*) which is itself exponentially decreasing with time (as may be the case for population reproduction, constrained growth, and other situations):

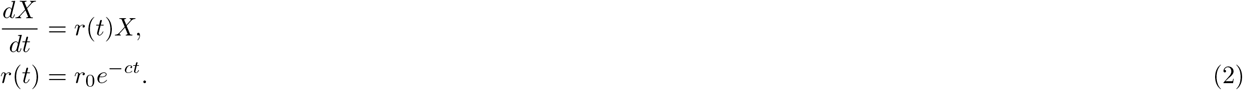

The solution to this is

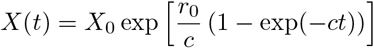

which we reparametrize as

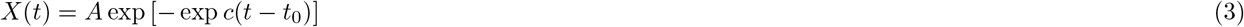

identifying *A* ≡ *X*_0_ exp(*r*_0_*/c*) and 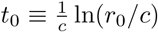 Tjørve and Tjørve [15] observe that this parametrization is more useful than others because of its “easy interpretable parameters”: *A* is an overall scale factor and the upper asymptote, *c* is the retardation rate of the growth rate *r* (eq 2), and *t*_0_ is a time offset (it represents the time at the inflection point of the curve).

For each ultrasound biometry measurement, we fit *t*_0_ and *c* over our population of fetuses but fit *A* individually for each fetus. Our data, and comparisons with published growth standards and tables, suggest that *t*_0_ and *c* can be treated as universal parameters, and almost all variation in normal fetal growth is explained by the scale parameter *A*. Similar to INTERGROWTH’s formulas for the mean and standard deviation of each biometry measurement, we compute a population mean *A* and its standard deviation.

### Quality of fit

The mean squared error (MSE) was computed for each participant using values derived from the equation and their corresponding real measurement data. Fetuses were ordered in ascending order by their mean square error, with the bottom 10% of data in each biometry classified as a bad fit for that biometry. Fetus for whom at least 2 out of 4 biometrics were bad fits were classified as overall bad fits (implying perhaps abnormal growth) and the rest as good fits or normal growth. By this criterion, every fetus was classified as either a “good fit” or a “bad fit”. To see whether bad fits are indicators of neonatal complications, the distribution of neonatal complications in the full set, the good fits, and the bad fits were analyzed. Significance was assessed via *p*-values using a hypergeometric test. Power analysis was performed by simulating 10,000 sets with the same size and proportion of bad fits, with the effect size that we actually observe; the power is the fraction of the sets where the null is (correctly) rejected by the hypergeometric test.

### Predicting birth weight of fetus

A predictive model was developed for the prediction of birth weight from ultrasound measurements taken at 24 weeks or earlier, or 35 weeks or earlier, using linear regression. Linear regression fits a linear model that makes a linear relationship between input features and target variables. The parameter regression coefficient and intercept are estimated by least square methods i.e it minimizes the sum of squared errors.

For each fetus, the parameter *A* was computed using measurements taken up until 24 weeks or up until 35 weeks. Biometries at delivery were predicted using the Gompertz function. These biometries, and their corresponding *A* were used as input parameters for birth weight prediction. Birth weight was predicted using the “leave one out” technique, where all data except the fetus under consideration are used for training and validation.

These predicted biometries and *A* for each fetus, and global *t*_0_ and *c*, were re-learned at each leave-one-out step. This ensures that there is no data leakage, i.e. at no point is the data on which the model is tested ever used in training the model. Models were implemented in python with the scikit-learn package. We compared our results with the INTERGROWTH (4), the four Hadlock (5) and Shepard (6) formulas for the estimated fetal weight (EFW) [21]. Hadlock 2 (using AC, FL, BPD) is presented here, and the other three in supporting fig S3 and supporting table S1.

INTERGROWTH (units: g, cm):

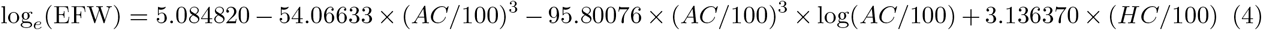

Hadlock 2 (units: g, cm):

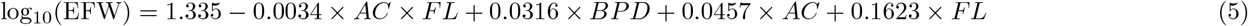

Shepard (units: kg, cm):

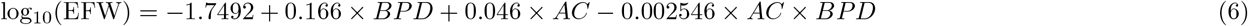

The model was evaluated by computing the mean absolute percentage error (MAPE) and root mean square error (RMSE) using predicted and actual birth weights, in all cases.

### Validation cohort

The validation cohort was obtained from South Indian Pregnancy Research Network (SIPNET), with data contributed from six hospitals in Chennai, Pondicherry, Hyderabad and Kochi, India. As with the Seethapathy cohort, the validation cohort was screened for subjects with at least three available sets of ultrasound biometry measurements, resulting in 365 subjects.

Birth weights were predicted with fetal biometries as input, using the machine learning model constructed on the Seethapathy cohort. The linear regression model was trained on the full Seethapathy cohort, and then predictions were made on the validation cohort.

## Results

### Gompertz parameters for Seethapathy data

The parameters in the Gompertz equation were fitted to the data, separately for each ultrasound measurement, *t*_0_ and *c* globally over all fetuses, and *A* individually for each fetus. Table 3 shows the resulting optimized values of these variables.

**Table 3.**
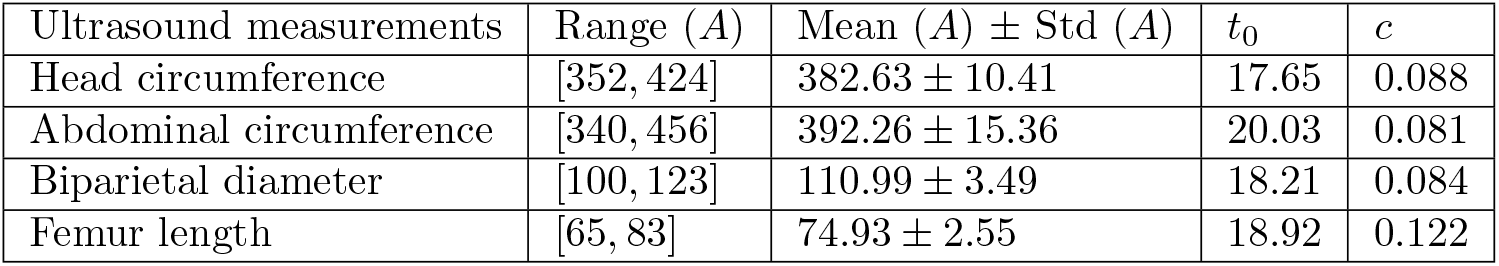
Gompertz parameters fitted to the Seethapathy cohort

*A* is an overall scale factor for each parameter, and its distribution over the individual fetuses is shown in figure 2A. Each fetus’ mean squared error (MSE) was determined by comparing the fitted function (with optimized *A*) with the original measurement. Figure 2B shows the distribution of the MSE. Figure 2C and D show, respectively, the five best and worst-fitting fetuses to the Gompertz curve.

### Neonatal complications are associated with deviations from Gompertz growth

We hypothesize that fetuses that do not fit well in their growth to the Gompertz curve could exhibit abnormalities in their growth patterns, with possible consequences for their health. We call the fetuses falling in the highest 10% mean squared error (MSE) to their Gompertz fit for an ultrasound measurement “bad fits” for that measurement. We further classify those fetuses that are bad fits in at least 2 out of 4 measurements (HC, AC, BPD, FL) as “overall bad fits” and the remainder as “overall good fits.”. Out of 817 fetuses in our cohort, 71 are “overall bad fits”. We find that neonatal complications are significantly associated with “overall bad fits” (p=0.007, hypergeometric test) as shown in figure 3A, with 31.0% of “overall bad fit” fetuses having neonatal complications versus 17.7% of “overall good fit” fetuses. Simulations suggest that for this effect size, the statistical power to detect a genuine effect with our cohort at *p* = 0.05 is 0.78.

Further, when we looked at the individual neonatal complications in both categories it was observed that SGA and LGA have a higher percentage (12.5% and 4.2% respectively) in the bad fit category as compared to the good fit category(3.4% and 1.4% respectively) figure 3B.

### Predicting final biometry and individual birth weight from early biometry

It is of clinical interest to predict the birth weight of an infant during gestation. We predict birth weight using, as input, our predicted ultrasound measurements at delivery and individual *A* values for each fetus (using the Gompertz formula) as described in Methods. Supporting figure S2 shows that our estimates of biometries at delivery are correlated with the actual birth weights.

Figure 4 shows birth weight predictions when we use available ultrasound data up until 24 weeks (green) and 35 weeks (blue), using linear regression, compared with the predictions of the INTERGROWTH, Shepard, and Hadlock 2 formulas. (The four Hadlock formulas are shown in supporting table S1 and their performance shown in supporting fig S3.

**Fig 4.**
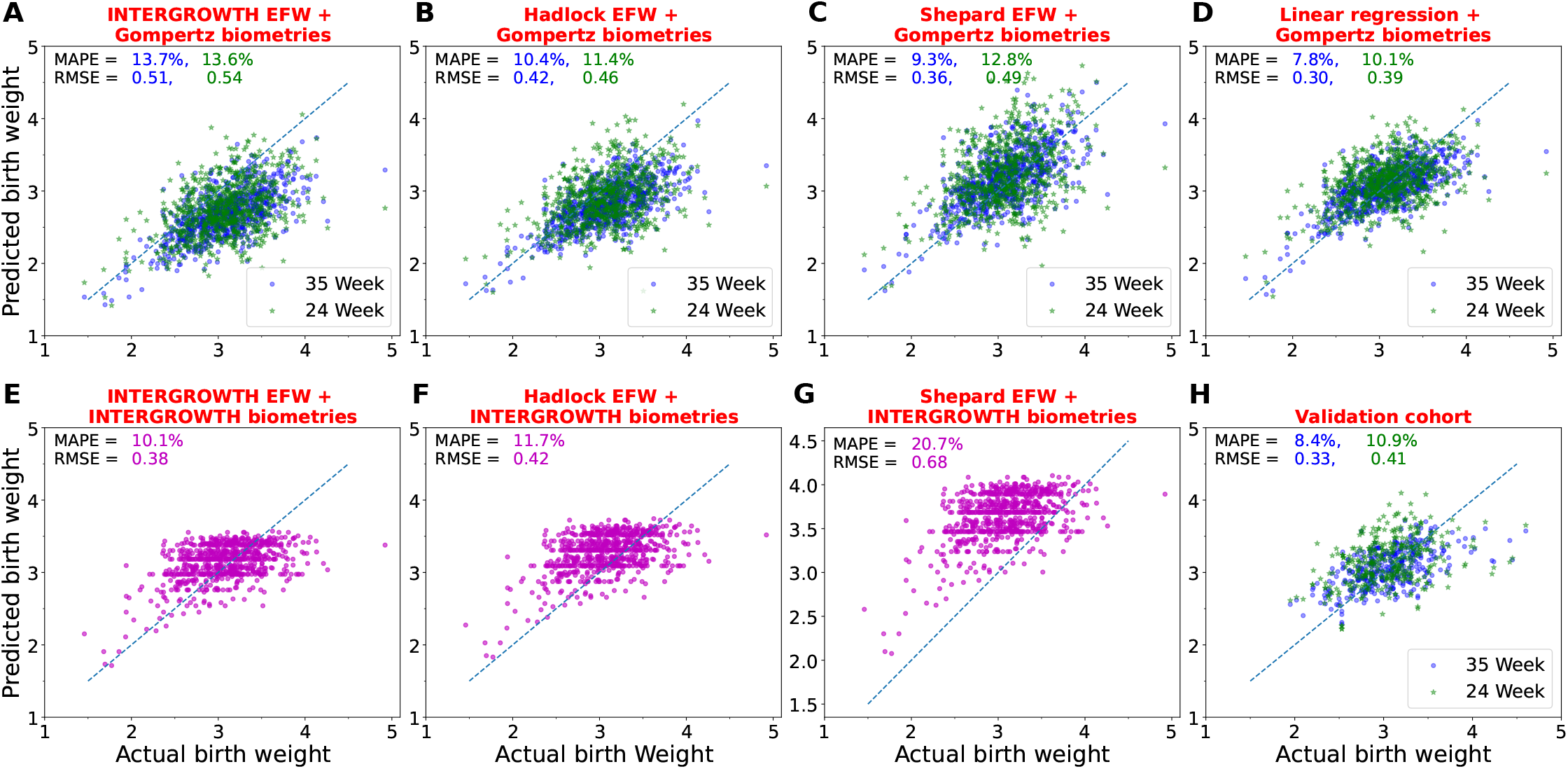
Birth weight prediction at delivery, using formulas from INTERGROWTH, Shepard, Hadlock, and linear regression. **A–C:** The final fetal biometries predicted by Gompertz expression, with *A* inferred from ultrasound scans up until 24 weeks (green) or 35 weeks (blue), were plugged in to the literature formulas to predict birth weight. **D:** Birth weight predicted by simple linear regression using Gompertz-predicted biometries. This was done in a leave-one-out manner to avoid data-leakage (Methods). **E–G:** Birth weight predicted using literature formulas with the INTERGROWTH mean biometry values for the gestational age at delivery. **H:** Birth weight prediction of the model trained on the full Seethapathy cohort, on an independent validation cohort, again with *A* calculated from 24 weeks (green) or 35 weeks (blue) scans.

Hadlock 2 performs best.) In all these formulas we used our predicted values of fetal biometries at term using the Gompertz equation and INTERGROWTH mean formula. Our individualized prediction, from early ultrasound parameters, clearly outperforms population-average formulas in predicting birth weight for individual fetuses. The MAPE for predictions made using all ultrasounds up to 35 weeks is about 7.8%. This information can be useful to clinicians making decisions on intervention. Figure 4 shows the biometries obtained from INTERGROWTH mean formula saturates after some time hence not able to predict the larger birth weight’s fetuses wheres Gompertz predicted biometries show a linear correlation with birth weight (supporting figure S2).

Equation 7, obtained using linear regression, can be used to estimate birth weight for fetuses, given estimates of final biometries and of the Gompertz scale parameters obtained from available ultrasound measurements.

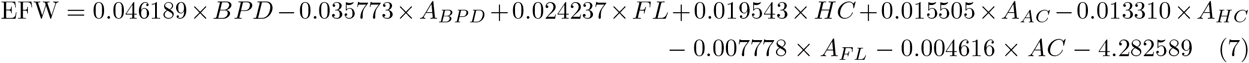

Marginal improvement was obtained using maternal parameters and LASSO regression (supporting figure S1).

Fig 5 shows, as a heatmap, the percentage of fetuses that fall within various prediction accuracies. In particular, with ultrasound data up until 35 weeks, our linear regression formula predicts 71.3% of birth weights within 10% accuracy. Shepard with Gompertz parameters (35 weeks) is next best at 63.8% cases predicted within 10% accuracy. Supporting figures S4 and S5 show the percentages under- and over-predicted by each method; it appears that our LR mathod is balanced, while INTERGROWTH and HADLOCK underpredict when using Gompertz biometry predictions, and Shepard overpredicts when using INTERGROWTH biometry estimates.

**Fig 5.**
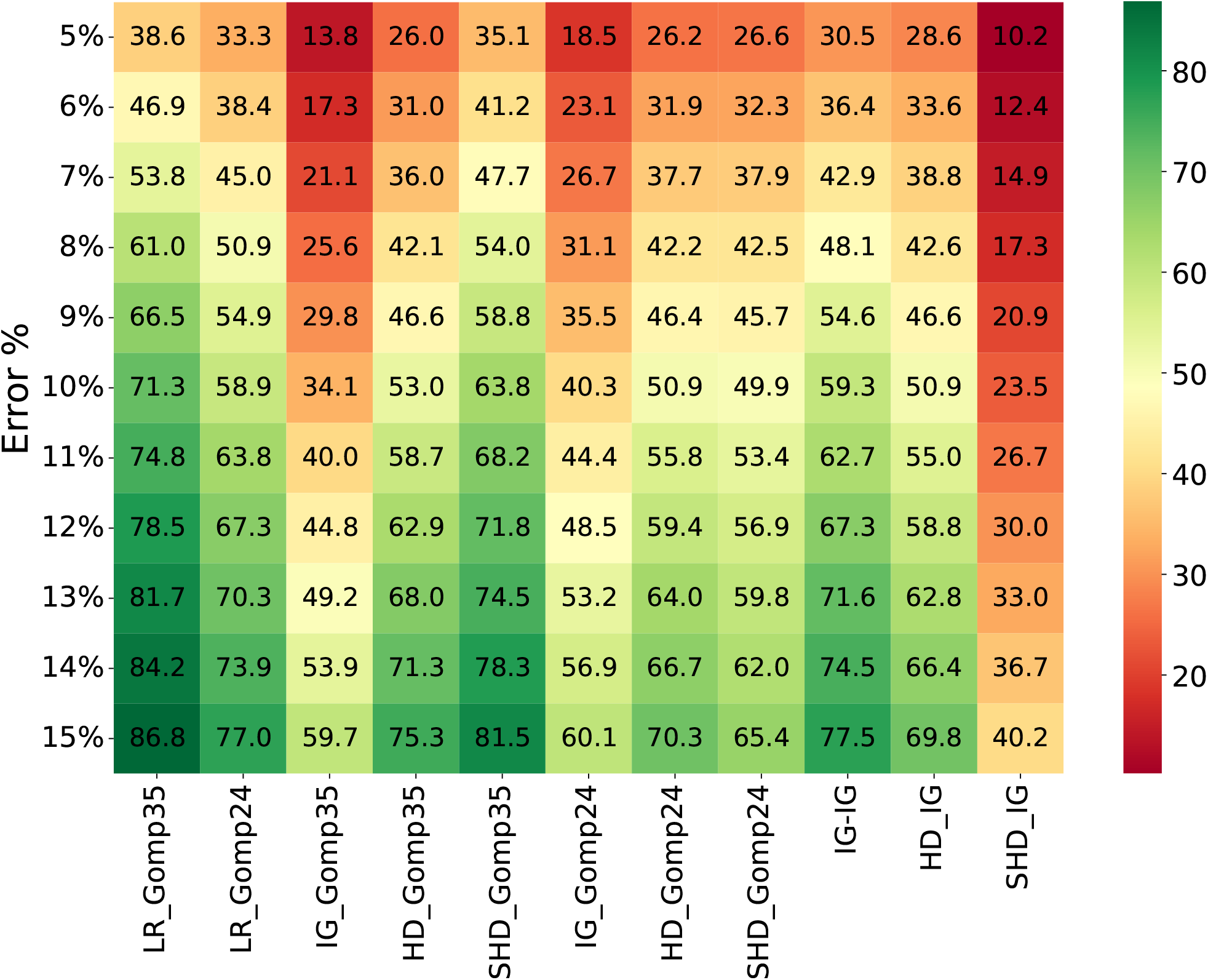
The percentage of fetuses whose birthweight is predicted within a given error rate, according to 11 methods. Here, LR=our linear regression formula, IG=INTERGROWTH, HD=Hadlock, SHD=Shepard, Gomp24=Gompertz prediction of final biometries using ultrasound data up until 24 weeks, Gomp35=Gompertz prediction of final biometries using ultrasound data up until 35 weeks. So, for LR Gomp35 and IG Gomp35, the Gompertz-predicted final biometries were used in our linear regression model and in the INTERGROWTH formula respectively. For IG IG, HD IG, SHD IG, the INTERGROWTH optimal fetal biometries at birth were used for prediction.

### A model trained on the Seethapathy cohort performs well in birth weight prediction on validation cohort

As described in Methods, we trained a model on the Seethapathy cohort and then predicted individual birth weights on the validation cohort. Using ultrasound data up until 24 weeks and up until 35 weeks, MAPE and RMSE on the validation cohort (fig. 6) are comparable with our predictions on the Seethapathy cohort.

**Fig 6.**
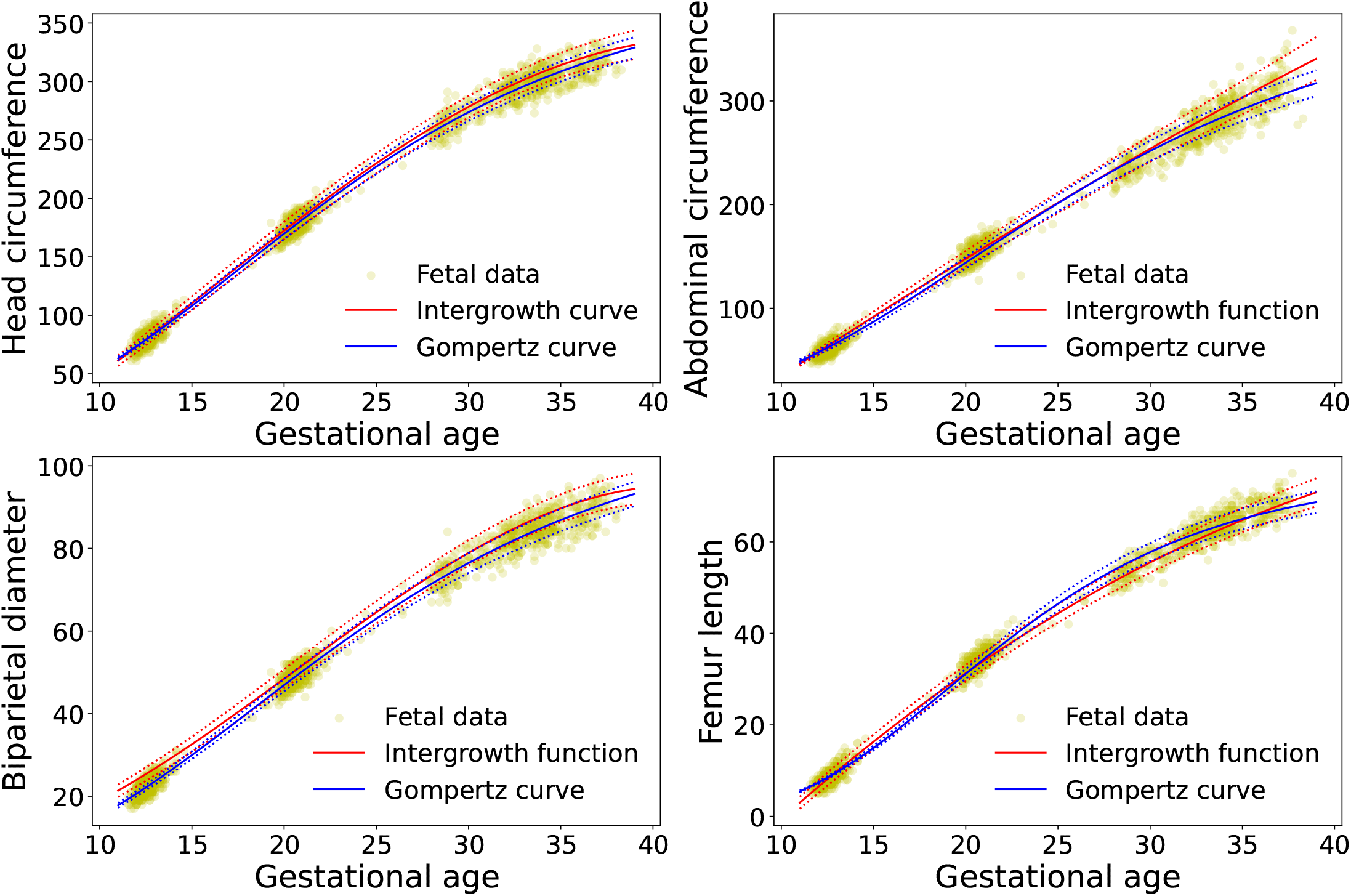
Comparison of INTERGROWTH growth curve with Gompertz curve. Actual fetal data (yellow) compared with Gompertz (blue) and INTERGROWTH (red); solid line = mean, dashed line = mean ± standard deviation; for Gompertz, we used the mean and standard deviations of *A* in our population.

### Framing fetal growth standards using the Gompertz equation

In recent years, the INTERGROWTH project [10], the NICHD, NIH, USA [7] and the WHO [5] have produced fetal growth curves for populations of healthy mothers with healthy lifestyles, and low-risk histories; and INTERGROWTH supplies equations as growth standards (table 2).

Fig 6 compares the fit of our fetal data with both the Gompertz and the INTERGROWTH curves. In this figure, the INTERGROWTH (red) mean curves are solid lines and mean ± standard deviations are dotted lines. For comparison, we have used the population mean *A* to plot the solid Gompertz curve (blue), and the population means ± standard deviation for *A* for dotted lines. Despite INTERGROWTH being a growth standard based on an entirely different cohort that was screened for numerous attributes relating to health (including minimum height and weight), socioeconomic status, and lifestyle, there is a remarkable agreement. There are some discrepancies: for biparietal diameter, the INTERGROWTH curves seem to slightly but observably overshoot our data.

In figure 7A we compare the INTERGROWTH mean and standard deviation curves for each parameter (black) with the Gompertz curve that fits best with that curve. Figure 7B does a similar comparison with the WHO fetal growth charts. (NICHD published separate charts according to ethnicity; we do not examine them here, but figures 3-5 in [8] compare their curves for AC, HC and FL with WHO and INTERGROWTH, suggesting small but significant ethnic differences.) The fit for INTERGROWTH was done in the 14-40 week window since that is the window where INTERGROWTH defines its standards. In the case of WHO, we observe a discrepancy in shape for AC and FL (and, slightly, for BPD), where growth seems to accelerate in the last weeks; we are unsure of the reason for this, so fit it in the window of 14-35 weeks (but the plot is over the full window of 14-40 weeks).

**Fig 7.**
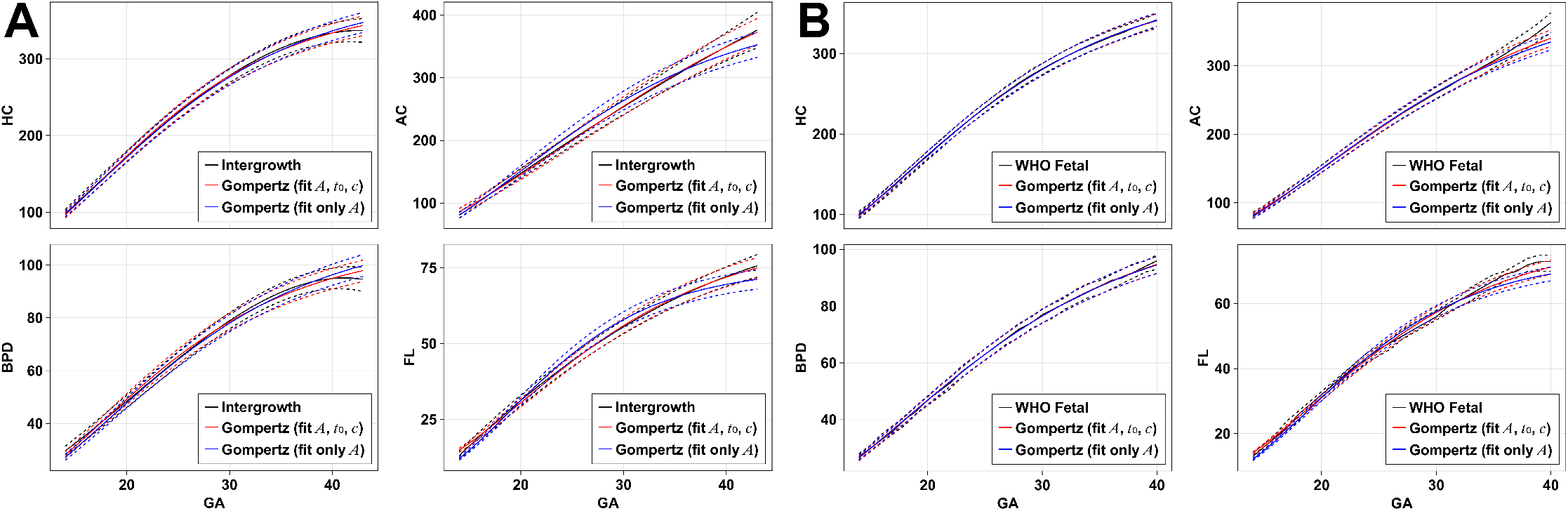
**A:** The INTERGROWTH growth standards for HC, AC, BPD, and FL (black); the Gompertz formula with *A, t*_0_ and *c* fitted to best match INTERGROWTH between 14-40 weeks (red); and the Gompertz formula with *t*_0_ and *c* obtained from our cohort (as in table 3) and only *A* fitted (blue). Solid lines are mean, dashed lines are mean ± standard deviation (INTERGROWTH) or *A* ± Δ*A* (Gompertz) where Δ*A* is chosen to best fit the INTERGROWTH dashed lines. **B:** A similar comparison with WHO data; here the dashed lines correspond to 25th and 75th percentiles for WHO, and the corresponding Δ*A* for Gompertz. The values of *A, t*_0_ and *c* for the red curves are listed in tables 4 and 5.

In all eight cases, in this window, the black (INTERGROWTH or WHO) and red (Gompertz) lines are essentially indistinguishable except, for some biometries, in the last few weeks.

**Table 4.**
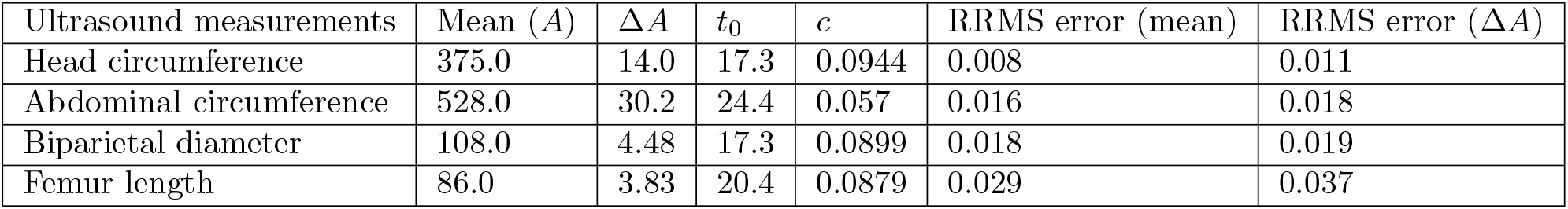
Gompertz parameters corresponding to INTERGROWTH growth standards (Δ*A* corresponds to standard deviation for INTERGROWTH)

| Ultrasound measurements | Mean ($A$) | $\Delta A$ | $t_0$ | $c$ | RRMS error (mean) | RRMS error ($\Delta A$) |
| --- | --- | --- | --- | --- | --- | --- |
| Head circumference | 375.0 | 14.0 | 17.3 | 0.0944 | 0.008 | 0.011 |
| Abdominal circumference | 528.0 | 30.2 | 24.4 | 0.057 | 0.016 | 0.018 |
| Biparietal diameter | 108.0 | 4.48 | 17.3 | 0.0899 | 0.018 | 0.019 |
| Femur length | 86.0 | 3.83 | 20.4 | 0.0879 | 0.029 | 0.037 |

**Table 5.** Gompertz parameters corresponding to WHO fetal growth curves (Δ*A* corresponds to 25th and 75th percentiles)

| Ultrasound measurements | Mean ($A$) | $\Delta A$ | $t_0$ | $c$ | RRMS error (mean) | RRMS error ($\Delta A$) |
| --- | --- | --- | --- | --- | --- | --- |
| Head circumference | 390.0 | 10.4 | 17.6 | 0.0893 | 0.005 | 0.008 |
| Abdominal circumference | 432.0 | 15.1 | 20.8 | 0.0746 | 0.004 | 0.008 |
| Biparietal diameter | 111.0 | 3.51 | 18.3 | 0.0842 | 0.007 | 0.009 |
| Femur length | 80.7 | 2.5 | 19.5 | 0.102 | 0.026 | 0.029 |

The corresponding parameters for mean *A*, standard deviation of *A, t*_0_ and *c* are listed in table 4 and 5. Also reported there are the relative root-mean-square difference from the standard, which, for INTERGROWTH, ranges from 1%–2% (except for FL which is a little higher) and, for WHO, is *<* 1% (again, except for FL).

Of interest, we note that in the case of HC and BPD in particular, the best-fitted values of *t*_0_ and *c* are very close to the values we obtained on the Seethapathy cohort (table 1), and the discrepancy is not large for AC and FL. The blue curves in figure 7 show the best Gompertz fit to the INTERGROWTH and WHO standards, using the values of *t*_0_ and *c* obtained from our cohort and only *A* fitted to the INTERGROWTH or WHO curves. While the the agreement is not as exact as between the red and black curves, it is still very good, suggesting that *t*_0_ and *c* can indeed be considered universal parameters for fetal growth, and the variation in fetal size is almost entirely described by the single parameter *A*.

The growth standard equations given by INTERGROWTH are hard to interpret in terms of a growth model, while WHO and NICHD do not supply equations. We suggest that the Gompertz growth model is a close fit for fetal growth, fitting the data to a better than 1% accuracy (WHO), better than 2% accuracy (INTERGROWTH), except for femur length where the error is slightly larger. We encourage future researchers to consider the Gompertz formula for describing fetal growth standards.

## Discussion

The Gompertz growth formula is a formula describing constrained growth, derived from an assumption that the growth rate is itself retarded at a constant rate *c*. It has two other parameters: *A*, which is an overall scale and *t*_0_, which represents the inflection time of growth. We demonstrate that this intuitive formula, used on the Seethapathy cohort, fits fetal biometry data with high accuracy. Almost all variation between fetuses can be described via the single scale parameter *A*, while *t*_0_ and *c* can be regarded as universal. This enables us to use the formula predictively, learning *A* from ultrasound data to predict final fetal dimensions. The simple linear regression model built on these predictions result in a *<* 8% MAPE of birth weight. This is remarkable considering that we use scan data done before 35 weeks. In contrast, previous studies cited in Introduction [20–22] used subjects who had an ultrasound scan within seven days before giving birth, but report similar or worse accuracies than us. Such late ultrasounds are not routinely done and the ability to predict final fetal biometries can be useful.

The previous work by Wosilait *et al*. [11, 16] applied this formula to fetal volume (or, almost equivalently, fetal weight). These are not directly measurable in utero. For the measurable ultrasound biometries, there exists one recent and comprehensive set of standards, the INTERGROWTH fetal growth standards [10], which are given as complex mathematical expressions. However, we show that the Gompertz formula, with appropriate parameters, is a close fit to the INTERGROWTH standards for HC, FL, AC, and BPD ultrasound measurements. We suggest that it can be a basis for future growth standards and, indeed, that INTERGROWTH standards can be reframed in terms of mean and standard deviation values for the single parameter *A*, treating *t*_0_ and *c* as universal.

The INTERGROWTH project has also published standards for estimated fetal weight (EFW) at birth [19], including a formula based on the expected values of the ultrasound biometries at birth. Previous such formulas include Hadlock1 [18] and Shepard [17]. In developing these formulas, a final fetal ultrasound was taken within one week of delivery. Ultrasound biometries are not typically taken so close to term, but these formulas can be used with personalized final biometries predicted by the Gompertz equation fitted to available ultrasound data, or with the “standard” biometry values as predicted by INTERGROWTH’s formulas. Our linear regression-based approach outperforms all these cases, with a slight further improvement seen on including maternal data.

We suggest that our algorithm of (a) estimating Gompertz growth parameters *A* for the fetus based on all available ultrasound, (b) using these to predict the parameters at term via the Gompertz formula, and then (c) using these parameters in our regression expression to estimate the fetal birth weight, is a significant improvement on existing formulas in the literature. In case a late-term ultrasound is in fact available, those measurements can be directly plugged into our formula.

Other than INTERGROWTH, there are also fetal weight standards published by WHO [5, 6] and NICHD [7, 8]. Both of these publish EFW curves using the Hadlock formula [18], as well as tables and charts for fetal biometries. While INTERGROWTH ignores country-specific or race-specific differences, describing the same standard for all healthy well-nourished fetuses, WHO and NICHD find inter-group differences despite no known environmental or socioeconomic factors. We find that the Gompertz formula with optimized parameters is a very good fit for the INTERGROWTH and WHO standards, with relative errors of less than 1% (WHO) and less than 2% (INTERGROWTH) except for FL which has a relative error of *<* 3% (both curves). Moreover, the values of *t*_0_ and *c* are close to the values obtained on the Seethapathy cohort, suggesting that these can indeed be treated as almost universal parameters and most fetal growth variability can be explained by the single parameter *A*.

As the authors of the WHO study [6] and NICHD study [8] have noted, the growth curves do seem to show significant variation by race and country. Future studies could explore the variation of the optimal *A*, and possibly also *t*_0_ and *c*, across such populations. Moreover, all three studies have chosen healthy populations of middle-to-high socioeconomic status, since the aim was to document ideal standards of fetal growth. But it is equally important to study fetal growth in undernourished populations, or in mothers with health complications. In such future studies, too, we believe the Gompertz formula will show its utility, but it will be particularly interesting to see if *c* (the rate of retardation of fetal growth) and *t*_0_ (the inflection time of growth) varies in such populations.

## Data Availability

All data produced in the present study are available online at https://github.com/chandranikumari/FetusGrowthModel

## Data and code availability

Anonymized data and the code that we used to plot graphs in this paper are available at https://github.com/chandranikumari/FetusGrowthModel

## Acknowledgements

The Gompertz function was suggested to us by Sitabhra Sinha in a comment on an early version of this work where we were using the Hill function. We also acknowledge useful discussions with Ponnusamy Saravanan and Suresh Seshadri. We thank Vanita Rajagopal and sonologists at Mediscan Systems, Chennai for performing the ultrasounds.

## Author contributions

**Conceptualization:** GIM,LN,UR,RS

**Data Curation:** CK, UR

**Formal Analysis:** CK, LN, RS

**Funding Acquisition:** GIM, LN, UR, RS

**Investigation:** CK, LN, UR, RS

**Methodology:** CK, GIM, LN, UR, RS

**Project Administration:** LN, UR, RS

**Resources:** LN, UR, RS

**Software:** CK, RS

**Supervision:** GIM, LN, UR, RS

**Validation:** CK, LN, UR, RS

**Visualization:** CK, LN, RS

**Writing – Original Draft Preparation:** CK, LN, RS

**Writing – Review & Editing:** CK, GIM, LN, UR, RS

