## Supporting information for "A predictive model of a growing fetus"

### Supporting Information for
### A predictive model of a growing fetus

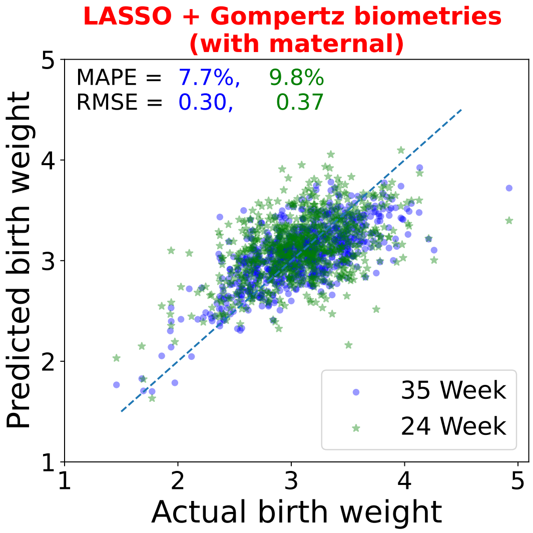


**Fig S1.** Birth weight prediction by using biometries at delivery predicted by the Gompertz function, $A$ calculated using scans at 24 weeks (green) and 35 weeks (blue) along with maternal parameters mentioned in Table 1, i.e., the first 11 rows. Here linear regression was used with the L1 penalty (also known as the LASSO model) to learn a model over all parameters. Again, leave-one-out was used similar to figure 4 in the main text to predict birth weight. We note a marginal improvement over the model that does not use maternal parameters.

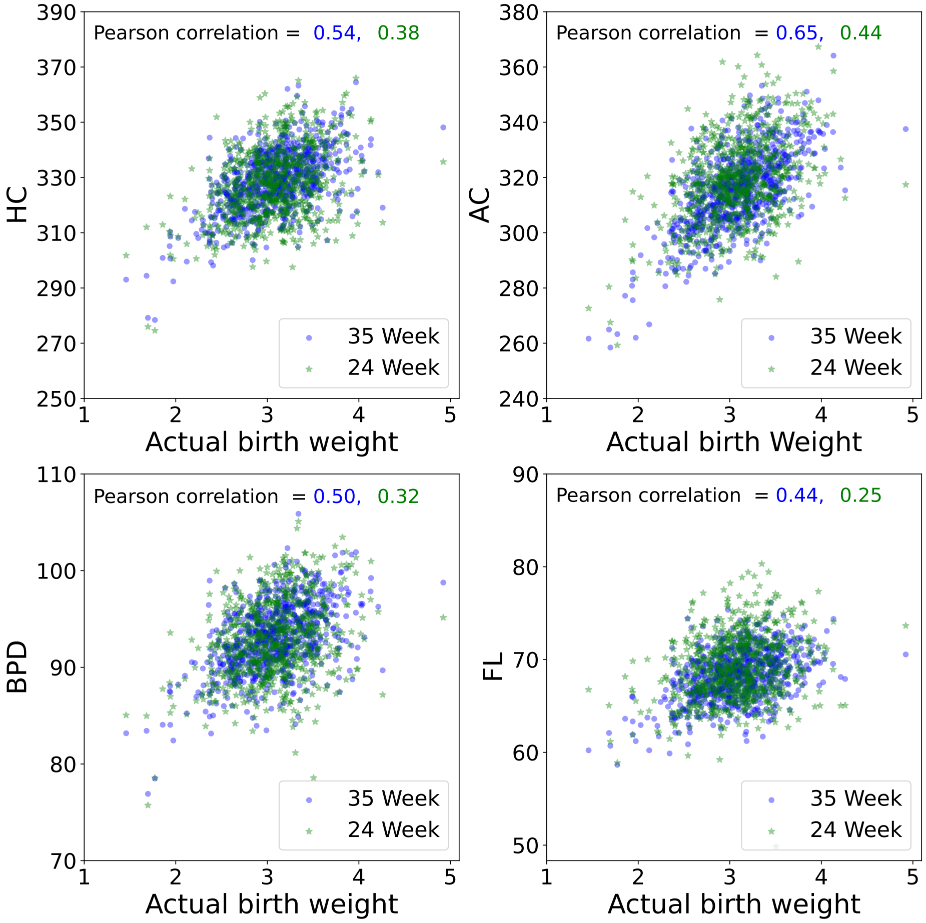


**Fig S2.** Scatter plot shows the correlation between the actual birth weight and the biometrics at delivery as predicted by the scans at 24 and 35 weeks (green and blue, respectively).

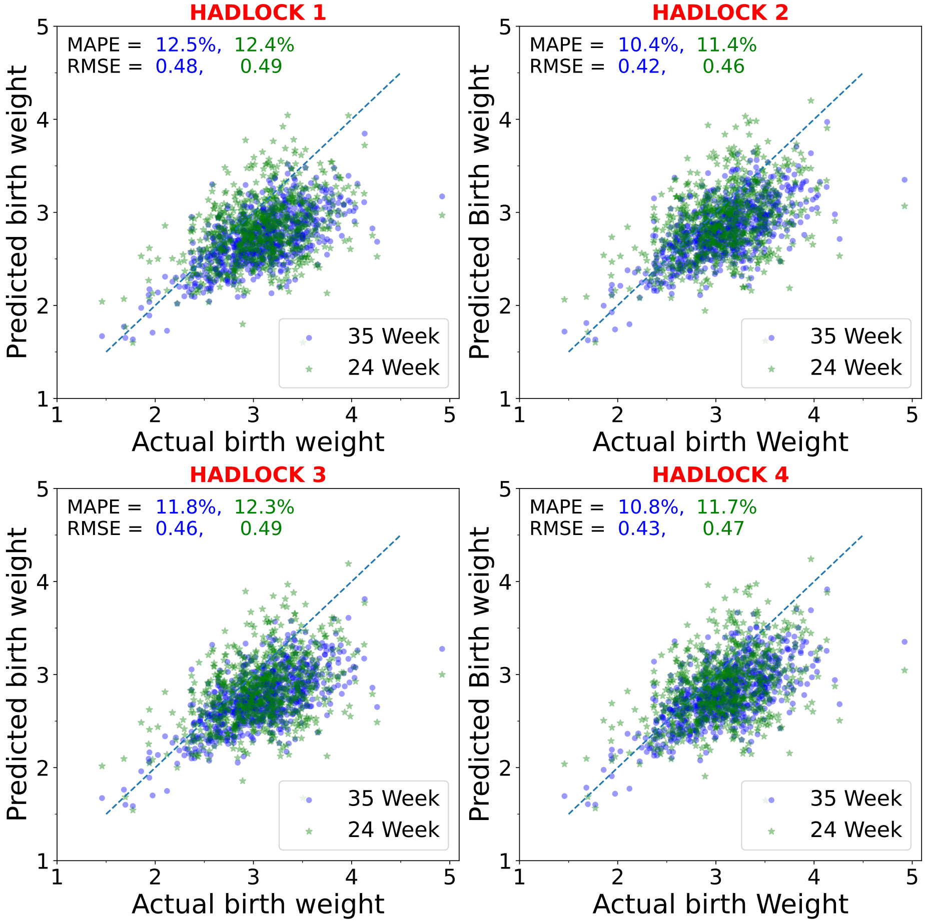


**Fig S3.** Birth weight prediction at delivery, using four different formulas from Hadlock. Biometries at delivery were predicted using the Gompertz function and $A$ was calculated using scans at 24 weeks (green) and 35 weeks (blue)

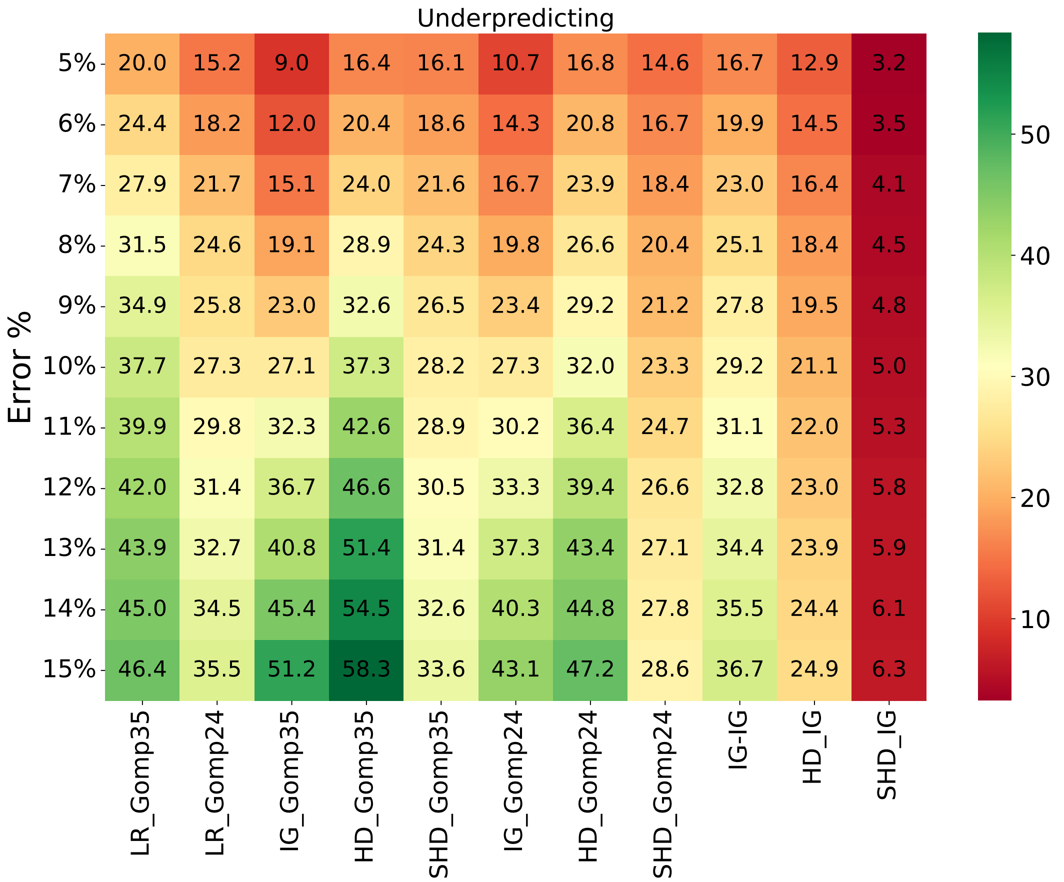


**Fig S4.** The percentage of fetuses whose birthweight is under-predicted within a given error rate, according to 11 methods.

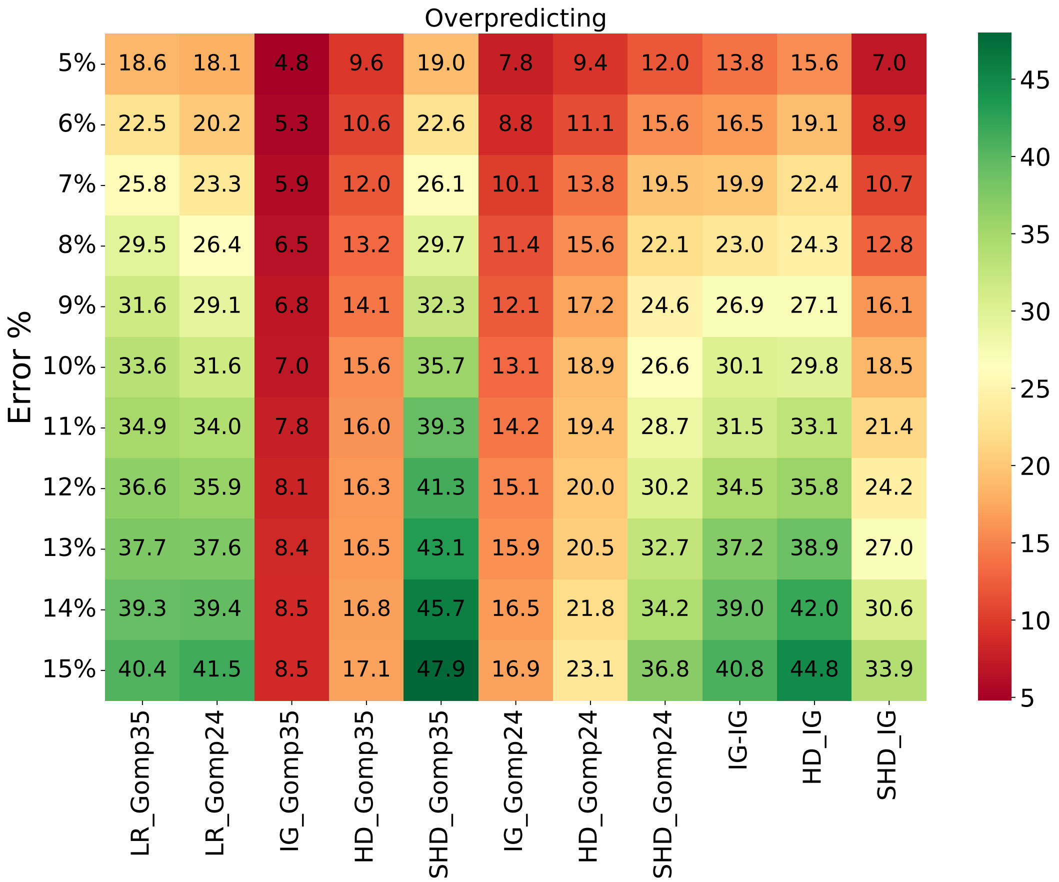


**Fig S5.** The percentage of fetuses whose birthweight is over-predicted within a given error rate, according to 11 methods.

**Table S1.** Four different HADLOCK regression formula for birth weight estimation

| | HADLOCK regression formula |
|---|---|
| HADLOCK 1 | $\log_{10}(\text{EFW}) = 1.304 + 0.05281 \times AC + 0.1938 \times FL - 0.004 \times AC \times FL$ |
| HADLOCK 2 | $\log_{10}(\text{EFW}) = 1.335 - 0.0034 \times AC \times FL + 0.0316 \times BPD + 0.0457 \times AC + 0.1623 \times FL$ |
| HADLOCK 3 | $\log_{10}(\text{EFW}) = 1.326 - 0.00326 \times AC \times FL + 0.0107 \times HC + 0.0438 \times AC + 0.158 \times FL$ |
| HADLOCK 4 | $\log_{10}(\text{EFW}) = 1.3596 - 0.00386 \times AC \times FL + 0.0064 \times HC + 0.00061 \times BPD \times AC + 0.0424 \times AC + 0.174 \times FL$ |
